# Tracking deaths from hitherto undetected infections can be an indicator of latent sars-cov-2 cases

**DOI:** 10.1101/2021.06.02.21258217

**Authors:** Yashaswini Mandayam Rangayyan, Sriram Kidambi, Mohan Raghavan

## Abstract

**Background:** With countries across the world facing repeated epidemic waves, it becomes critical to monitor, mitigate and prevent subsequent waves. Common indicators like active case numbers can flatter to deceive in the presence of systemic inefficiencies like insufficient testing or contact tracing. Test positivity rates are sensitive to testing strategies and cannot estimate the extent of undetected cases. Reproductive numbers estimated from logarithms of new incidences are inaccurate in dynamic scenarios and not sensitive enough to capture changes in efficiencies. Systemic fatigue results in lower testing, inefficient tracing and quarantining thereby precipitating the onset of the epidemic wave.

**Methods:** We propose a novel indicator for detecting the slippage of test-trace efficiency based on the numbers of deaths/hospitalizations resulting from known and hitherto unknown infections. This can also be used to forecast an epidemic wave that is advanced or exacerbated due to drop in efficiency.

**Results:** Using a modified SEIRD epidemic simulator we show that (i) Ratio of deaths/hospitalizations from an undetected infection to total deaths converges to a measure of systemic test-trace inefficiency. (ii) This index forecasts the slippage in efficiency earlier than other known metrics. (iii) Mitigation triggered by this index helps reduce peak active caseload and eventual deaths.

**Conclusions:** Deaths/hospitalizations accurately track the systemic inefficiencies and detect latent cases. Based on these results we make a strong case that administrations use this metric in the ensemble of indicators. Further hospitals may need to be mandated to distinctly register deaths/hospitalizations due to previously undetected infections.

**Key Messages:**

- Deaths or Hospitalizations are unmissable events in an epidemic and this paper proposes a metric *D*_*ratio*_ based on these numbers to monitor the inefficiencies in test-track-trace performance.
- The ratio of deaths(or hospitalizations) resulting from undetected infections to total deaths (or hospitalizations) detect the onset of laxity in regulations earlier than other conventional metrics like daily increase in active cases, daily deaths or even reproductive number estimates.
- Mitigation by tracking the *D*_*ratio*_ reduces or truncates the epidemic wave intensity or delays it sufficiently.

## 1. Introduction

Compartmental epidemiology models like SIS,SIR,SEIR and SEIRD model the effects of changing dynamics in an epidemic in the form of transitions through a sequence of compartments [1, 2, 3, 4]. These transitions are a result of changes in number of individuals in each state as the individual subjects move from susceptible to getting exposed, infected and finally recover or die. The reproductive number R0 in an epidemic is the mean number of infections an infected person can cause in a fully susceptible population during the lifetime of a person’s infection [5]. R0 indicates the attack rate or spread of infection with values greater than 1 indicating build-up and values less than 1 indicating fade out of the disease [6, 5, 7]. This metric is useful in gauging the risk of an epidemic [8] and when estimated in real time can determine the effectiveness of intervention strategies like lock downs and isolation that change the transmission dynamics [6, 8].

Modified SEIRD model [9] uses the epidemic simulator as tool for administration and governance. It uses compartments in two parallel tracks representing the quarantined and unquarantined population which makes it relevant in the days of COVID 19 pandemic. This model demonstrated the utility of using simulators for predicting the number of infections in both the quarantined and unquarantined arms of the model in the normal course and in response to a variety of administrative actions, thereby estimating the effectiveness of administration reforms. These models empower the policy makers and health workers to understand the model parameters better and take necessary and timely interventions. It is well known that testing helps flatten the curve while undetected and latent cases spur on a full blown epidemic. The epidemic simulations [9] helped to quantify this effect as a measure of inefficiency.

Reasons for undetected cases are many. People who are asymptomatic or with limited access to healthcare (forming a large part of the population) might go undetected easily and hence the number of confirmed, recovered and deceased cases that are reported everyday can be an underestimate of the true figures [10, 11, 12]. Further insufficient testing, contact tracing or quarantining may contribute to this inefficiency. Amidst all the chaos of asymptotic cases, under-reporting etc, death is an unmissable event in an epidemic. The number of deaths in an epidemic provides a realistic picture of the extent of infection still latent in the population [13, 14]. While it is intuitively recognized that detection of infection postmortem or at first presentation at intensive care units are an indicator of the gravity of situation, this has never been quantified or studied as a measure or indicator of relevant quantities. Current reporting practices do not capture ‘infections detected postmortem’ or ‘previously undetected infections presenting at ICU’ separately from the detected infections. We propose that these metrics are vital indications of the health of a community or society and can be an early indicator of an emerging epidemic wave.

Using the epidemic simulator described in [9], we demonstrate the utility of such metrics in early detection of an epidemic wave that is hastened or exacerbated by inefficiency [9] on account of insufficient testing, tracing or containment measures. We use the symmetry in the two arms of the model [9] along with the fact that death of either kind is unmissable, to estimate the inefficiency (or latent spread) of an epidemic. We show here that recording the number of ‘postmortem detection of infection’ as a proportion of total deaths can be a good metric. The ratio of ‘previously undetected infections at hospitalization’ to total hospitalizations would also serve a similar purpose. We hereby make a case for modifying the Standard Operating Procedures to make this book keeping possible.

## 2. Methods

The Modified SEIRD model described in [9] uses two symmetric, parallel arms to represent the quarantined and free populations. The transition rates from one compartment to another are proportional to the extent of contact tracing and self-reporting in each of the respective compartments. These transition rates are described in Figure 1 [9].

**Figure 1:**
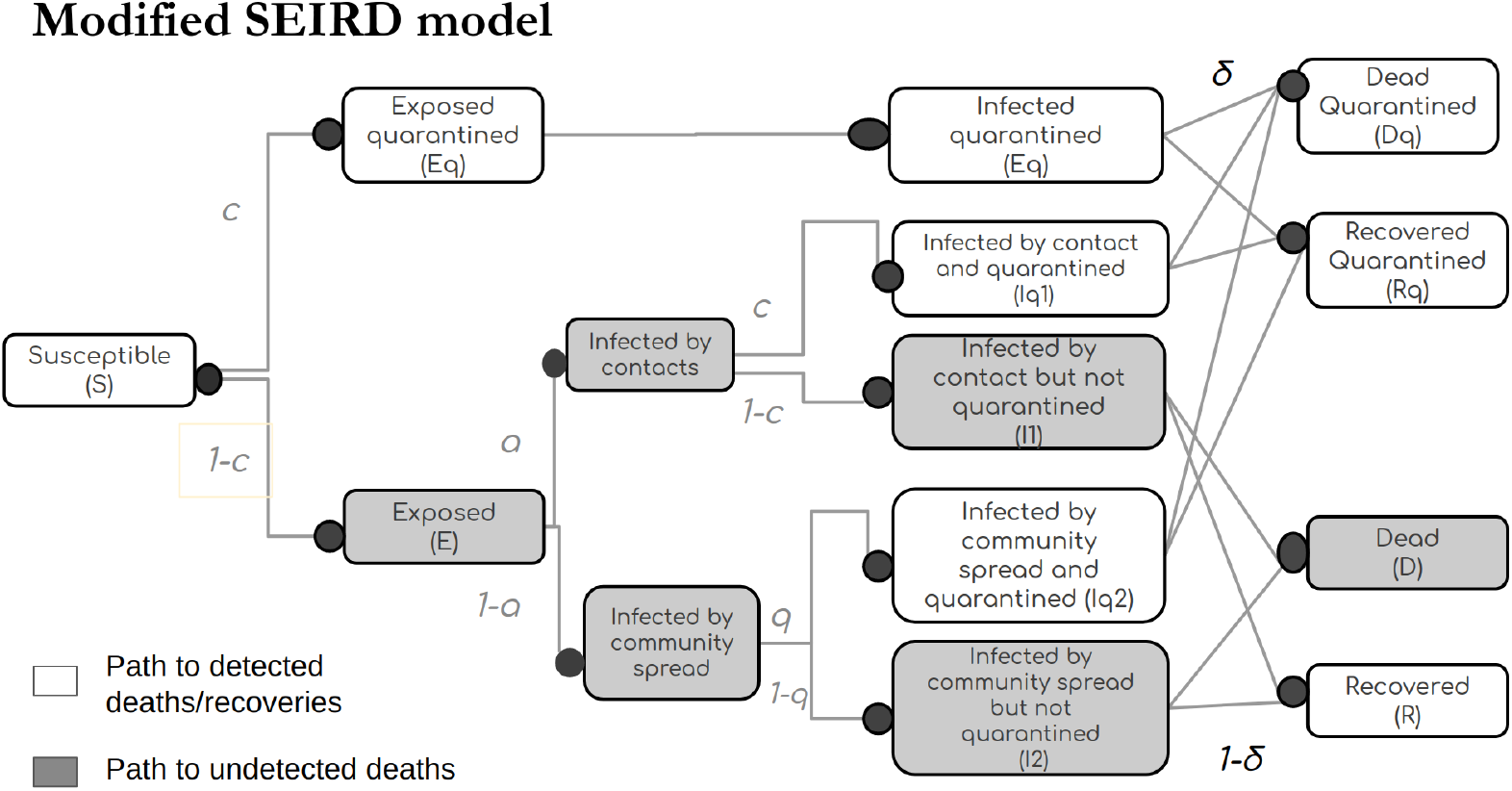
Figure Adapted from [9] describes the fraction of population flowing through each compartment in the parallel arms of the Modified SEIRD model.

These transitions provide a metric called the intervention inefficiency that is interpreted as the ineffectiveness of interventions; or the fraction of infections that are undetected. The inefficiency being a fraction in [0,1] is the complement of efficiency [9].

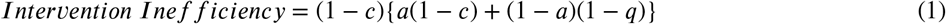

where,

a -The fraction of infection spread through contacts,

q -Fraction of infections detected through random testing and self reporting,

c -Fraction of infections detected through contact tracing.

It is observed that c and q play a significant role in altering the inefficiency.

The ratio of unreported cases to total number of cases can be captured in any of the compartments of the model at I or beyond, to measure the inefficiency. Since death is an unmissable event in an epidemic, the data for it is more easily and accurately available than in any other compartment. We thus hypothesize that calculating the ratio of unreported deaths to total deaths is a good indicator of inefficiency. Alternatively if the compartments D and Dq are replaced by H and Hq for hospitalisation of hitherto undetected infection and hospitalization of a known infection, the inefficiency can also be interpreted as ratio of ‘previously undetected infections at hospitalization’ to total hospitalizations.

This paper uses a measure *D*_*ratio*_ to describe the ratio of unreported deaths to the total number of deaths obtained from the deceased compartment.

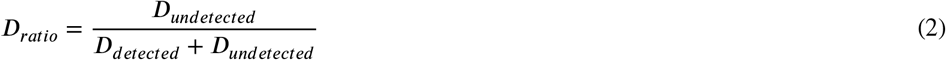

While the structure of the compartment model itself guarantees that the *D*_*ratio*_ will asymptotically converge to the inefficiency when parameters of the model are constant, we wish to explore the utility of *D*_*ratio*_ in scenarios where inefficiency is varying dynamically. In order to test our hypothesis, we probe the relationship between the *D*_*ratio*_ and the intervention inefficiency and their growth trends in a simulated epidemic with changing intervention inefficiencies. In particular we are interested in exploring the scenarios where a drop in efficiencies due to systemic fatigue in a prolonged epidemic can exacerbate a wave in infections.

### Convergence of *D*_*ratio*_ to true inefficiency and lag in convergence

The experiments are simulated by seeding the model with an initial set of parameters as described in the Table 1. This is done to ensure the model simulates a realistic number of cases in each of the compartments and it also furnishes an identical backdrop for all the experiments.

**Table 1.** The model parameters used for simulations

| Parameter | Value |
| --- | --- |
| $inf_t$ - mean infection time | 9 |
| $lat_t$ - mean latency time | 7 |
| $in_{phi}$ - rate of population inflow 21 | 5 |
| pl - probability of infection in the population inflow | 0.1 |
| Number of days predicted by the simulator | 125 |
| Number of days simulated before the 1st infection | 50 |
| Population size | 10 000 000 |
| a - fraction of infections through contact | 0.3 |
| $\beta$ | 0.35 |

Interventions are introduced at two instances of time t1 and t2 having corresponding intervention inefficiencies of Inefficiency1 and Inefficiency2. Each of these Inefficiencies are varied from 0 to 1 in steps 0.05, and their corresponding time series of *D*_*ratios*_ are plotted, thereby exploring the ability of a variety of change patterns.

If the interventions are ineffective, the *D*_*ratios*_ increase and similarly when the interventions contain the spread of infection more effectively the *D*_*ratios*_ tend to decrease. Hence the Intervention Inefficiency becomes the true value of the *D*_*ratios*_. The lag with which the *D*_*ratios*_ converge with the prevalent inefficiency value is a good indicator of the utility of this metric. Hence the experiments run with various intervention inefficiencies are noted for observations. The time to convergence is defined as

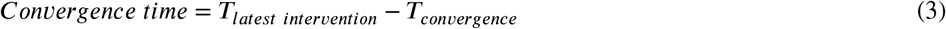

where,

*T*_*latest intervention*_ represents the the day on which the latest intervention was made,

*T*_*convergence*_ is the first day in a block of five days where the *D*_*ratio*_ is within ±5% of true inefficiency.

### The effect of window length on *D*_*ratio*_ curves

The *D*_*ratios*_ can be calculated in multiple ways; by summing over the entire epidemic period, or over windows of several days. The experiment was run over different window periods where the *D*_*ratios*_ were smoothed over the window length to omit the baggage of deaths that occurred much earlier in time. This led to the term *D*_*smooth ratios*_ defined as :

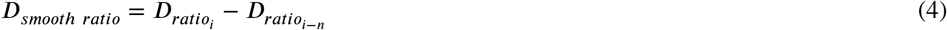

where,

i represents the ‘i’th day,

n represents the number of days chosen in the window period.

### Comparison of using *D*_*ratios*_ and active cases for corrective action

Laxity in contact tracing and testing are common during a long ongoing epidemic due to systemic fatigue or ignored clusters. This leads to increase in inefficiency and case numbers are subdued or increase only mildly when the epidemic starts to rise. Thus it blindsides the system to a wave building up. We simulate such a case with inefficiency1 = 0.5 and an inefficiency2 rising to 0.7 which results in a wave. A response to this situation results in a decrease in inefficiency to 0.3. The timing of the response and its effect on the impending wave are explored. In this scenario we also explore the resultant rise in *D*_*ratios*_ and active cases and their relative utility in predicting and mitigating the impending latent wave.

## 3. Results

### The ratio of unreported deaths to total deaths converge with their true value of inefficiency

It may observed from Figure 2 that the *D*_*ratio*_ tracks the true value of inefficiency as the inefficiency changes dynamically. The metric tracks the true inefficiency on both the increasing and decreasing excursions. While the time to converge varies, the changes in *D*_*ratio*_ follow a largely exponential trajectory. As a result, changes in the metric of interest are large at the onset of change and becomes progressively slower. This suggests that the *D*_*ratio*_ can be a good metric for detecting sudden changes in underlying inefficiency.

**Figure 2:**
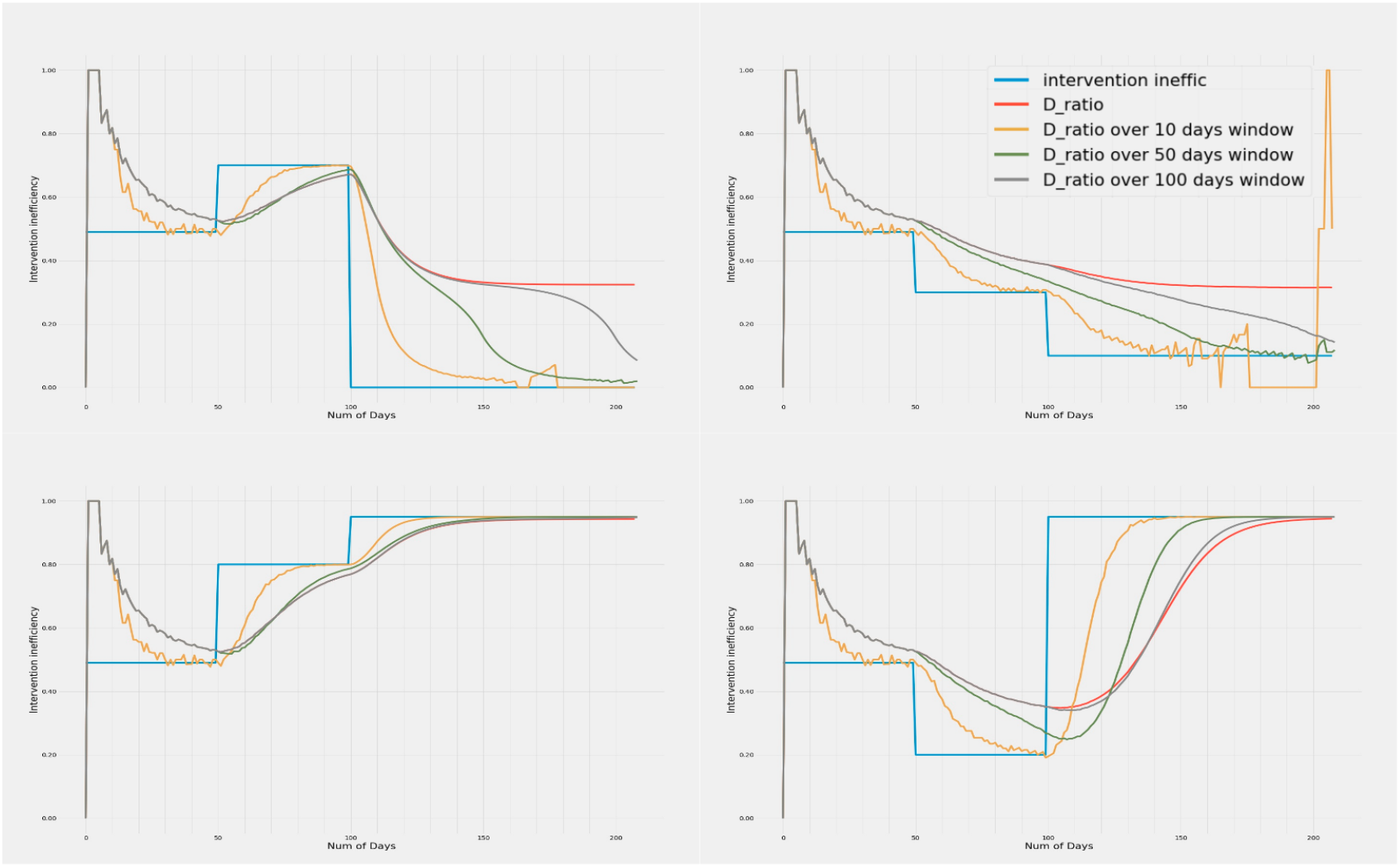
Variation of Intervention inefficiency and *D*_*smooth ratio*_ with time. The 4 plots show the trends in *D*_*ratio*_ over window periods of 10, 50 and 100 days when the inefficiencies are increased and decreased in different temporal orders. Metrics calculated with all window sizes show high day to day variation when the death numbers in either arm is very low. However, metrics with smaller window sizes are more prone to this daily variation. It is observed that *D*_*ratio*_ when smoothed over a small window of 10 days, converges faster but has a lot of variance due to daily fluctuations in numbers. Whereas *D*_*ratio*_ over a 100 days window length is smoother, but takes longer to make inferences due to slower convergence. Although sensitive to noise, 10 day window period gives the fastest convergence followed by 50 and 100 day periods.

### *D*_*ratio*_ metric with varying window sizes and their tracking performance

It may be seen that the *D*_*ratio*_ calculated over the entire epidemic period suffers from memory effects and is unable to converge properly to the true inefficiency. Computing *D*_*ratio*_ over smaller windows results in faster convergence to the true inefficiency. The smaller windows also show larger gradients early on, hence are probably more useful in detecting a change in underlying inefficiency. However this involves some trade-off as with smaller time windows, the metric can become noisy due to daily variations, especially when the absolute number of deaths is very low due to very low inefficiency. The larger windows give smoother convergence and better estimates of underlying inefficiency when the number of absolute deaths in either arm is low. An appropriate window size (or an ensemble thereof) helps achieve a balance between reliability of inefficiency estimates with a faster response time in changing and reforming administrative policies.

### Convergence time to true inefficiency is directly proportional to differences in inefficiencies

The heatmap in Figure 3 shows the relationship between magnitude of change in inefficiency and the number of days for *D*_*smooth ratio*_ over a 50 day window to converge with the latest inefficiency. The bright diagonal band shows that convergence is the fastest when the difference between the inefficiencies are small and increase with difference between inefficiencies. This diagonal is flanked on both sides by a series of symmetric bands that progressively widen with increase in the inefficiencies. The broader bands of fast convergence in the bottom-right indicates that the metric performs progressively better and is advantageous in higher inefficiencies when the situation most warrants it. The convergences are slower when absolute number of deaths are low in either arm (detected or undetected) as seen when either of the inefficiencies is small. Such situations result in high variance of *D*_*smooth ratios*_ beyond the 5% margins used to define convergence in our definition.

**Figure 3:**
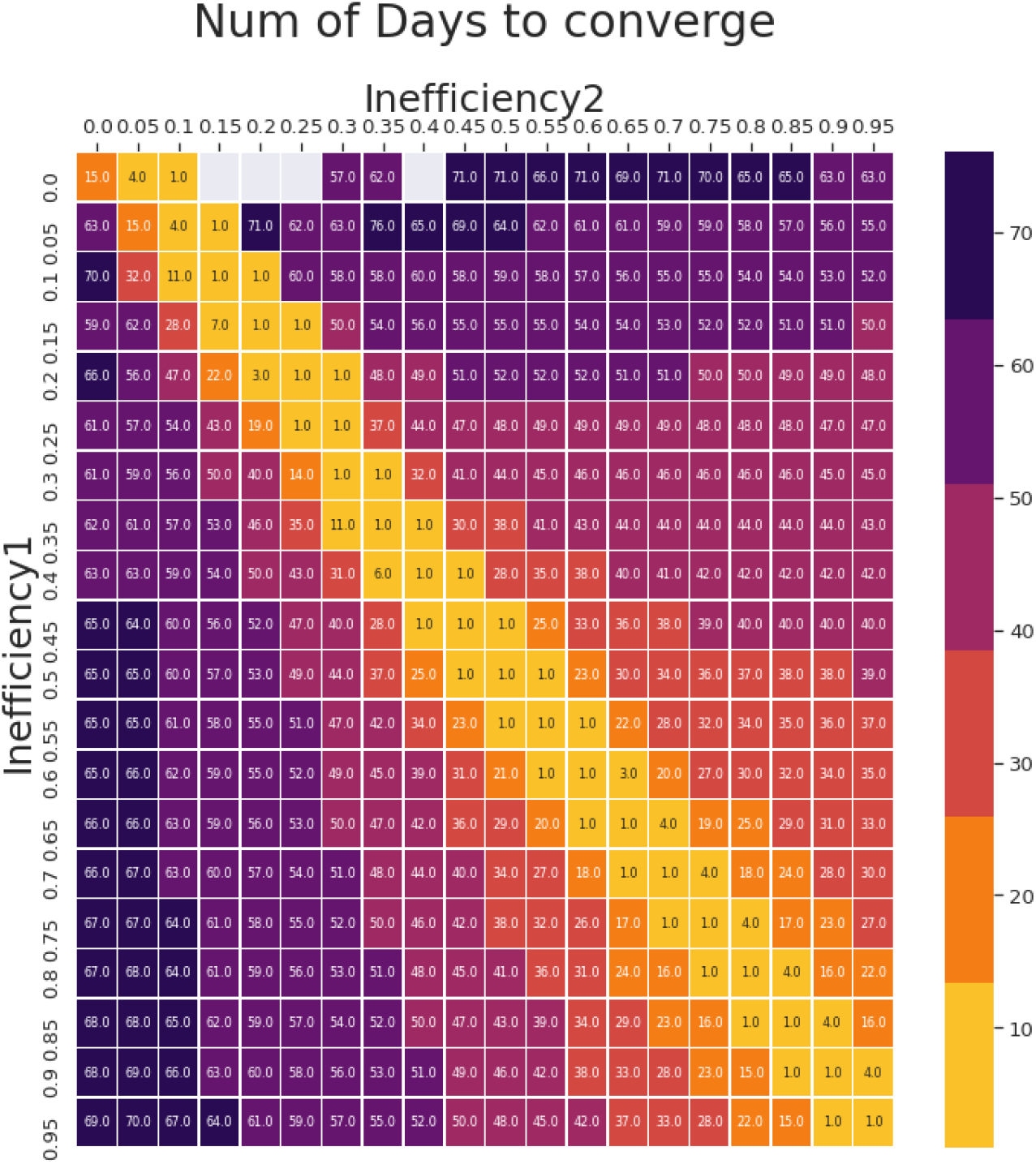
Correlation between Intervention *Inefficiency*_1_,*Inefficiency*_2_ when *D*_*smooth ratio*_ is calculated over a 50 day window and the number of days for *D*_*smooth ratio*_ to converge with *Inefficiency*_2_ is plotted on a heatmap.The days to converge is the least on the diagonal and is largely symmetric. Transitions in the range of inefficiencies 0.2 through 1 converge within a period of 60 days and the range from 0.4 through 1 within 50 days.The empty(grey) cells indicate it takes more number of days to converge than the period of simulation or the cases are extremely low to show up in the *D*_*smooth ratio*_. In general it may be seen that in the high inefficiency range where it is critical to react fast, the convergences are faster, thus emphasizing the utility of this metric.

### Smoothed *D*_*ratio*_ can detect onset of laxity earlier than other metrics

Figure 4 compares the responses of metrics such as daily increase in active cases and daily deaths with the *D*_*ratio*_ calculated over a ten day window. It may be noticed that the change in inefficiency causes an increase in *R* value from 1.57 to 2.20 thereby precipitating a wave. However responding in the wake of conventional metrics like daily increase in active cases or log changes in active cases are only visible after a time lag as the lower efficiencies result in a large fraction of cases to be undetected. If the same change in *R*_*t*_ had occurred by means of increased */J* instead of an increased inefficiency, the daily increase in active cases would have increased faster, with a possibility of early detection. When inefficiencies increase due to decreased testing or contact tracing, the daily increase in active cases is rather subdued in the short term, masking the impending wave. Although deaths cannot go unnoticed, rise in deaths can take even longer to show up sufficiently to raise an alarm. However, the *D*_*ratio*_ captures the imbalance between the detected and undetected arms and hence indicates the changes underneath. The daily change in the *D*_*ratio*_ can be an indicator of the noise in the estimation of *D*_*ratio*_. While full convergence of *D*_*ratio*_ to the true inefficiency may take time, it is seen from Figure 4 that within a span of about 20 days it converges to the new inefficiency and at 10 days already shows clear indications of rise in inefficiency as evidenced by sudden high increase and moderate variance.

**Figure 4:**
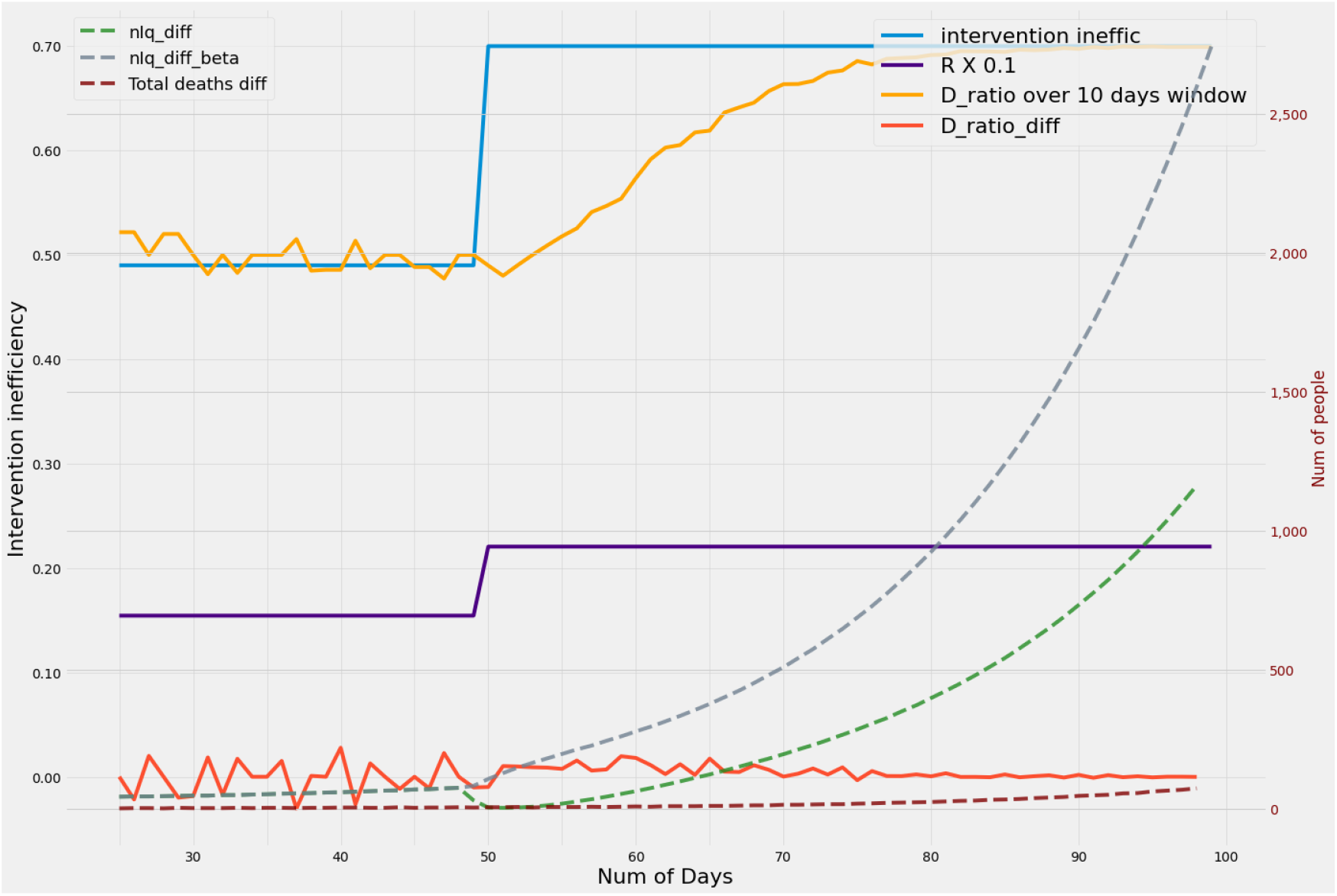
The graph considers the rise in active cases due to two scenarios, both of which result in identical changes in reproduction number R. When the rise in R is due to an increase in */J*, the active case increase yields a sharper curve giving a true indication of underlying cases. However, when the the increase in R is due to increase in test-track-trace inefficiency, the rise in active cases are slower and in fact even decrease over the short term as the increasing cases are missed due to high inefficiency. A noticeable increase in active cases can only be seen past 30-40 days after the change. However, the increase in *D*_*ratio*_ is already evident in 10 days from the day inefficiency increases and stabilizes within a period of 20 days. Complete convergence as defined in Figure 4 comes around 35 days. It may thus be seen that the maximum convergence times of *D*_*ratio*_ metrics are comparable to the early rises in active case load based metrics. Thus windowed *D*_*ratio*_ based metrics are ideally suited as triggers for initiating mitigation.

### Mitigation by tracking the *D*_*ratio*_ reduces or truncates the epidemic wave intensity or delays it sufficiently

As the number of active cases in a pandemic remains low for a long period of time, it is natural for some laxity to creep in the regulations and protocols followed. This results in an increase in the inefficiency as indicated in Figure 5(a) thereby precipitating or advancing an impending latent wave. Intervention resulting in decreased inefficiency can be triggered by one or more indicators. The interventions triggered after observing the conventional metrics like significant rise in the number of active cases would easily cost 60 days of valuable time. The earliest response monitoring the situation aggressively would also take no less than 40 days (Figures 5(b) and 5(c)]. Despite reducing the deaths and active cases,both these scenarios [Figures 5(b) and 5(c)] leave very little time to prepare or react resulting in quite a number of infections and deaths. Intervention based on sudden deviations of the metric *D*_*ratio*_ plays a seminal part here in averting the wave or pushing the wave much farther ahead in time as indicated in Figure 5(d), where the inefficiency is lowered based on the increase in *D*_*ratio*_. As seen in Figure 4, *D*_*ratio*_ can detect the true inefficiency in about 20-30 days with certainty as witnessed by very low daily variations in *D*_*ratio*_. The metric can clearly forewarn although with with slightly lower certainty (moderate daily variation) within 10 days. Using *D*_*ratio*_ the maximum time to track the inefficiency transition is given by the heatmap in Figure 3 and the earliest response time is about a half or third of it as observed in Figure 4. The benefits of reacting based on this metric is demonstrated in Figure 5(d). The peak active case load and death are reduced significantly. The early warning also provides the administrative bodies sufficient time to plan the interventions, resources and logistics needed to mitigate or tide over it.

**Figure 5:**
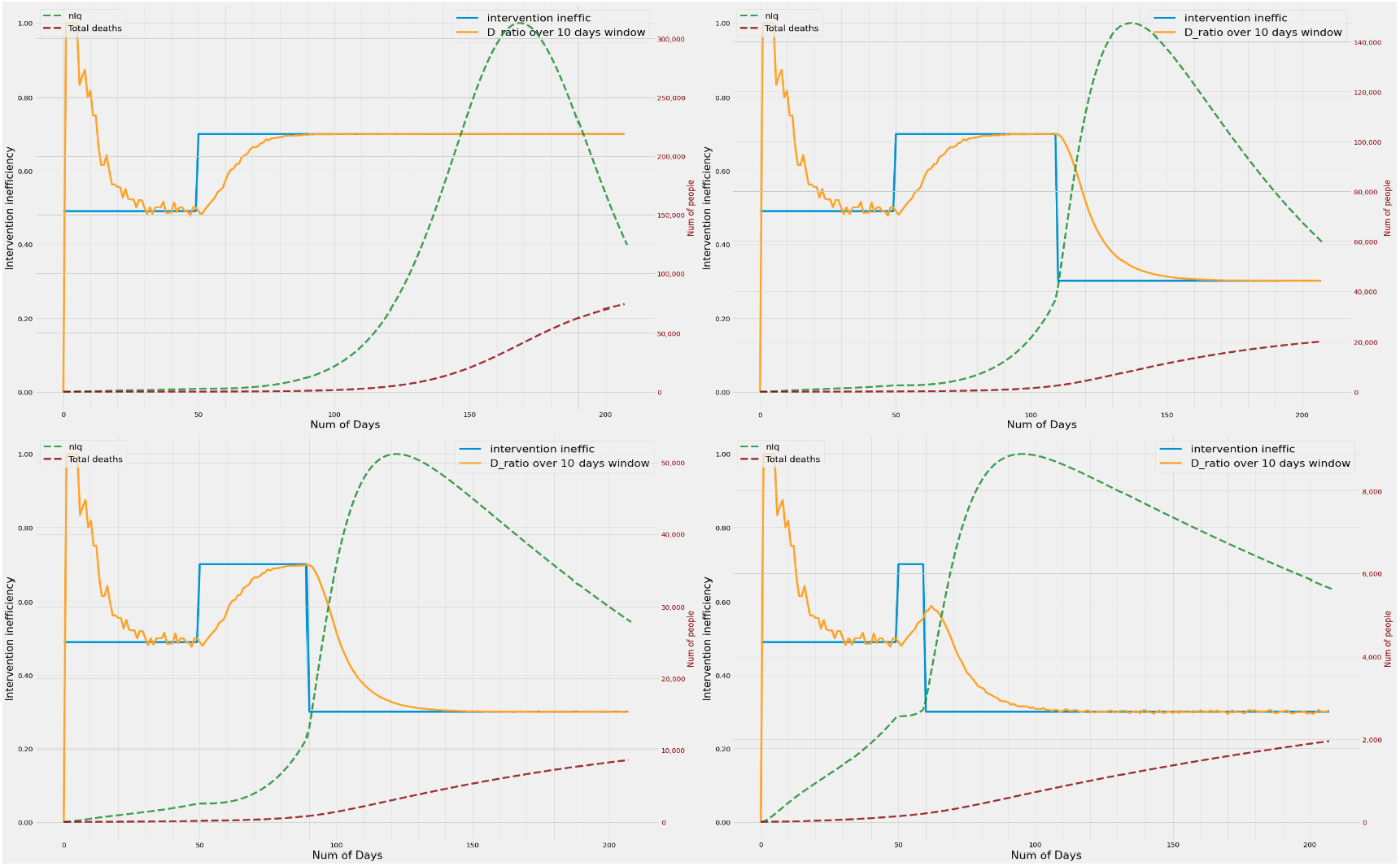
Left to right: **(a)** Inefficiency increases, but no counter measures are taken. Result is a peak active case load of 300 000 in a 10 million population and causing close to 75 000 deaths. **(b)** Inefficiency is lowered when the daily active cases significantly increase above 40,000. This measure reduces the active cases by nearly 50% and deaths by 75%. **(c)** By monitoring rise in active cases closely, lowering the inefficiency when they rise above a smaller threshold of 15 000 active cases daily, the peak and deaths both reduce considerably. **(d)** Instead when the inefficiency is lowered as the *D*_*ratio*_ begins to increase, the peak active cases and deaths are averted.

## 4. Discussions

Constantly monitoring an epidemic is extremely crucial as even small laxity in test-track-trace efficiencies has great potential to precipitate a wave. This situation can be analysed with the help of parameters like reproductive number, daily increase in active cases, test positive ratios etc. But these parameters are largely dependant on testing strategies and the nature of sampling, whereas monitoring deaths are unmissable events in an epidemic and are capable of explaining the underlying situation quantitatively. This paper proposes a metric based on death reports(or equivalently hospitalizations reported) to monitor the inefficiency in test-track-trace performance. We show that *D*_*ratio*_ being deaths resulting from hitherto undetected infections as a fraction of all deaths is sensitive to the changes in the inefficiencies prevalent in the system and reflects these changes quicker than any other conventional indicators. Our work also highlights the potential reduction in the peak active case load and deaths when mitigation measures are triggered by the metric *D*_*ratio*_. The proposed metric and inferences drawn, if employed as part of standard reporting procedures can enable the government to stay better equipped to predict a loss in efficiency and a resulting wave. The benefits can accrue by way of stronger mitigation and prevention of the wave at best or better preparation of hospital infrastructure (beds, ventilators, test kits etc) at the least.

Generalizing this method, the inefficiency thus defined could have been obtained from any of the compartments in the Modified SEIRD model [9] by taking the corresponding numbers from the two parallel arms for reported and unreported cases respectively. As deaths are usually reported at hospitals or government bodies, adequate data can be collected which is not guaranteed in other compartments of the model (namely S,E,I,R). Further these *D*_*detected*_ and *D*_*undetected*_ can be used interchangeably as *H*_*detected*_ and *H*_*undetected*_ representing the hospitalized cases or cases requiring intensive care with no loss of generality. *H*_*detected*_ would in this case mean hospitalization of a patient already diagnosed with the infection and *H*_*undetected*_ would mean the hospitalization of a patient due to severe symptoms but diagnosed with the infection post hospitalization. This extrapolation of results helps in arriving at the analysis faster and more importantly without risking a death.

This model assumes that the rates of death are the same in both the arms - detected and undetected. But in reality it is plausible that the undetected arm could have a lower probability of I->D transition than the detected arm. These different rates would show up as a scaling factor of *D*_*ratio*_. However, even in such scenarios the gradient of the rise in *D*_*ratio*_ compared to the previous epoch would still give indication of the impending wave earlier than the other indicators used currently.

In light of the above results, we strongly propose that policy makers and healthcare administrators consider the inclusion of *D*_*ratio*_ metric as part of their decision making framework. Since it is likely that hospitals already have this data, implementing this proposal would only demand certain changes in the book keeping of already available data, posing minimal overheads. By closely monitoring the trends in this metric it is possible to detect the changes in the laxity in regulations and take corrective measures much earlier and also gain ample time to work on the strategies and procurement of required resources to overcome the wave.

## Data Availability

All data and simulation related code are made available in the ver4.7 branch of the git repository : https://gitlab.com/yashaswinimr1/trackarona.git

## CRediT authorship contribution statement

**Yashaswini Mandayam Rangayyan:** Conceptualization of study, Methodology, Coding, Analysis of results, Manuscript preparation. **Sriram Kidambi:** Coding. **Mohan Raghavan:** Conceptualization of study, Analysis of Results, Manuscript preparation.

